# Performance of Vision-Language Models for Zero-Shot Lung Nodule Detection on Chest Radiographs

**DOI:** 10.64898/2026.05.31.26354565

**Authors:** Mizuho Nishio, Hidetoshi Matsuo, Takaaki Matsunaga, Koji Fujimoto, Nicolas Deperrois, Farhad Nooralahzadeh, Thomas Frauenfelder, Michael Krauthammer, Takamichi Murakami

## Abstract

**Background and Objectives:** The ability of vision-language models (VLMs) to detect lung nodules on chest radiographs remains uncertain. This retrospective study aimed to compare the zero-shot performances of six VLMs for lung nodule detection using data from the Japanese Society of Radiological Technology (JSRT) chest radiograph database.

**Methods:** A total of 247 chest radiographs from the JSRT database (154 with nodules and 93 without) were preprocessed and evaluated using six VLMs: RadVLM, gpt-4o-mini, Qwen3-VL-8B-Instruct, MedGemma-4b-it, LLaVA-Rad, and CheXpert Plus Model. Each model was tested using a zero-shot setting. The text outputs were binarized into nodule-present or nodule-absent labels by consensus between the two radiologists. Sensitivity, specificity, accuracy, precision, and F1 scores were calculated. Pairwise differences in sensitivity, specificity, and accuracy were assessed using McNemar’s test with Holm correction.

**Results:** The overall performance was limited across all models. RadVLM achieved the highest accuracy (44.5%, 110/247) with perfect specificity (100.0%, 93/93) and precision (100.0%); however, its sensitivity was low (11.0%, 17/154). LLaVA-Rad showed the highest sensitivity (27.3%, 42/154) and F1 score (37.7%), but lower specificity (71.0%, 66/93). MedGemma-4b-it achieved 100.0% specificity, with a sensitivity of only 5.2% (8/154). Grade-specific analysis showed that detection rates were highest for obvious nodules and remained limited for subtle nodules. Pairwise analyses revealed significant differences in sensitivity and specificity for the selected model pairs, particularly between RadVLM and LLaVA-Rad.

**Conclusion:** Current VLMs show limited zero-shot generalizability for lung nodule detection in the JSRT database, with marked trade-offs between sensitivity and specificity. Their near-term value may lie more in radiologist-assisted workflows than in stand-alone detection.

**Clinical Impact:** Current VLMs should not be used as stand-alone tools for lung nodule detection on chest radiographs because of their limited sensitivity and substantial model-dependent trade-offs. However, their high-specificity outputs in some models and higher-sensitivity behavior in others suggest potential roles in radiologist-assisted workflows, such as report drafting and second-reader support.

## Introduction

Lung nodules on chest radiographs (CXRs), clinically important findings, are among the most frequently overlooked abnormalities. They are often small, and their visibility can be reduced by overlap with the ribs, cardiac silhouette, hilar structures, and other anatomic shadows; therefore, detection rates may vary with the radiologist’s experience and attentional allocation [1,2].

Consequently, high-quality chest radiograph databases play an important role as resources for the rigorous evaluation of both human readers and computer-aided diagnostic algorithms. The Japanese Society of Radiological Technology (JSRT) database is a widely used benchmark for evaluating lung nodule detection [3].

Vision language models (VLMs) are multimodal models that jointly process images and text. Recent research includes domain-adapted biomedical and radiology VLMs in addition to general-purpose VLMs [4]. In chest radiography, these models have been explored for report generation, abnormality classification, visual grounding, and workflow support. RadVLM is a representative example of a radiology-oriented VLM [5].

However, major challenges remain in evaluating VLM performance for radiology report generation. Previous studies have assessed the agreement between natural-language reference reports and text generated by VLMs. However, in this framework, the diversity of expression, verbosity, and differences in clinically nonessential wording influence the evaluation results. Therefore, the conventional evaluation of a text generation task does not necessarily reflect the medical validity or practical performance of a specific clinical task [6].

Lung nodule detection provides a clear framework for evaluating this issue. Reducing VLM outputs to a binary classification of the presence or absence of a lung nodule lowers the ambiguity in evaluating free-text outputs, enabling clinically meaningful comparisons. Under this framework, the responses from each model can be judged according to uniform criteria and evaluated using conventional diagnostic imaging metrics, such as sensitivity, specificity, and accuracy.

Furthermore, evaluation under zero-shot conditions is important for determining the extent to which each VLM exhibits generalizable visual understanding and medical knowledge without additional training or task-specific optimization. From a practical radiology standpoint, evaluating VLMs on a high-quality external benchmark—distinct from typical training and assessment datasets—offers especially valuable insights [7]. However, a systematic zero-shot comparison of multiple current VLMs, including domain-specific and general-purpose models, on a standardized radiological benchmark has not been performed.

Therefore, this study aimed to evaluate the zero-shot performance and generalizability of multiple VLMs for lung nodule detection in the JSRT database using a binary classification framework.

## Methods

This study used the JSRT database; hence, the institutional review board approval and the requirement for informed consent were waived. A zero-shot evaluation is defined as an inference without task-specific fine-tuning, calibration, threshold optimization, or training on the JSRT dataset.

### Dataset

The dataset analyzed comprised 247 CXR images: 154 with lung nodules (100 malignant and 54 benign) and 93 without. Representative chest radiographs are shown in Figure 1. Based on the paper by Shiraishi et al. [3], the cases were selected from 14 medical institutions, and the CXR images were digitized in 12-bit grayscale. In the images with nodules, the nodules were categorized into five groups based on subtlety. In the original study [3], receiver operating characteristic (ROC) analysis was performed using the interpretations of 20 radiologists. In the present study, the accompanying JSRT labels of “nodule present” or “nodule absent” were used as the binary ground truth for each image.

**Fig. 1.**
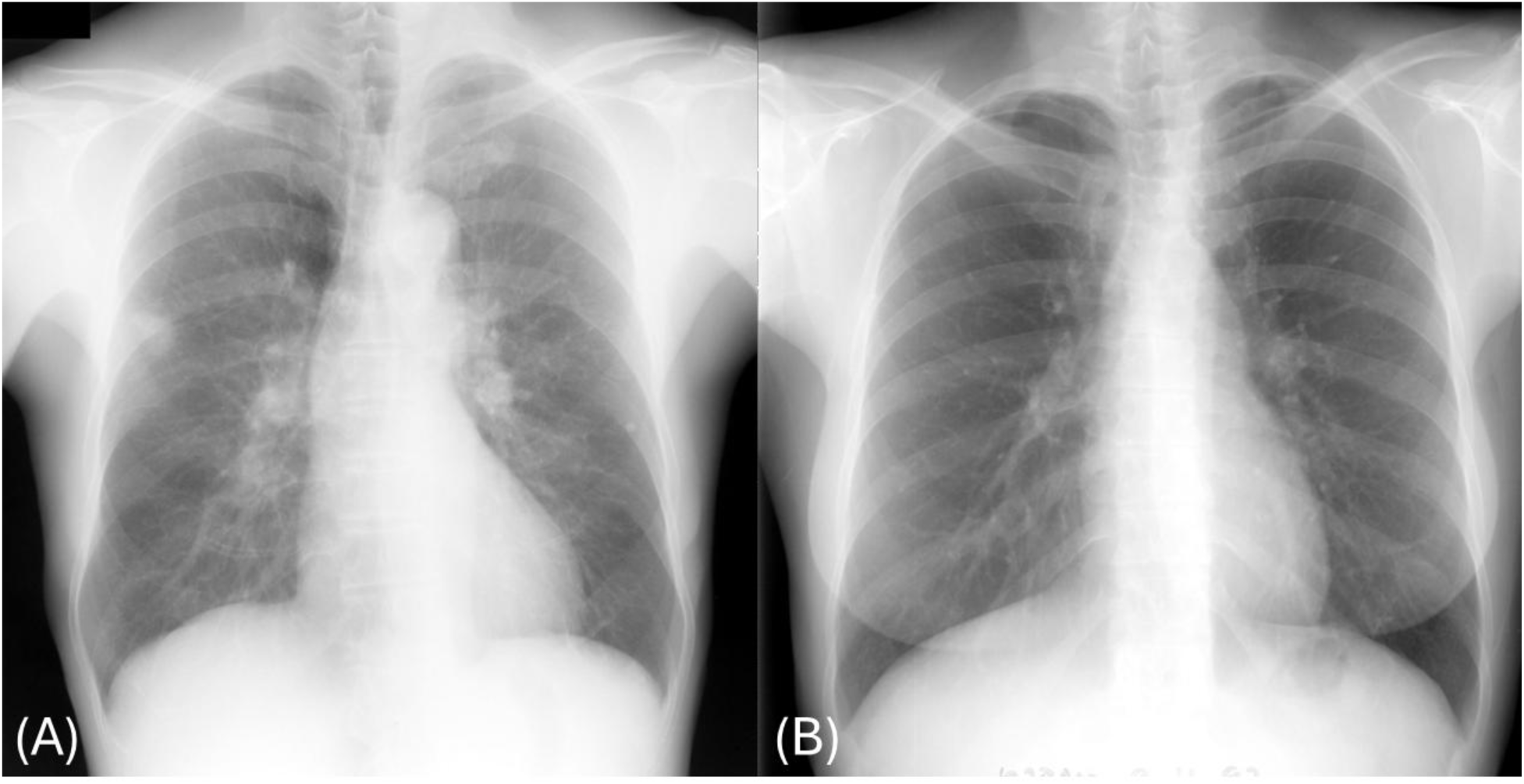
Representative chest radiographs from the JSRT database. Chest radiograph of (A) the right lung nodule; (B) no lung nodules. Abbreviation: JSRT, Japanese Society of Radiological Technology; CXR, chest radiograph.

### Image Preprocessing and VLM

Figure 2 and Table 1 show the image-processing workflow and the six VLMs evaluated in this study, respectively. The original JSRT CXR .img files, stored as 16-bit images with 12-bit grayscale data, were read and, when necessary, subjected to endian conversion to obtain 2048 × 2048-pixel images [3,8]. For each image, linear contrast correction based on the 2^nd^ to 98^th^ percentiles was applied, the image was converted to 8-bit depth, and saved in JPEG format as an inverted grayscale image. No cropping of the images was performed. The six VLMs were as follows:

- RadVLM [5]
- gpt-4o-mini (commercial service of OpenAI)
- Qwen3-VL-8B-Instruct [9]
- MedGemma-4b-it [10]
- LLaVA-Rad [11]
- CheXpert Plus Model [12]

**Fig. 2.**
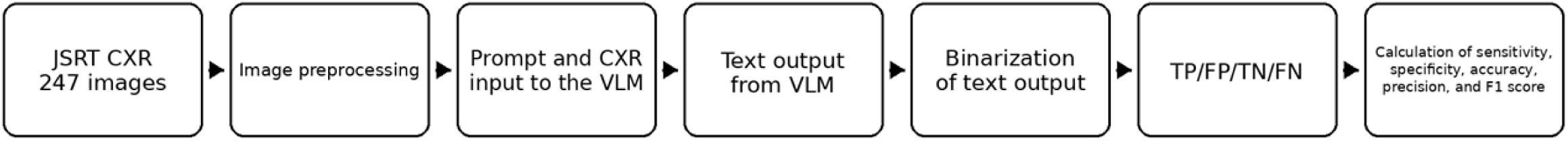
Workflow for image preprocessing and zero-shot evaluation of chest radiographs using VLMs. Abbreviations: CXR, chest radiograph; VLM, vision-language model; TP, true positive; FP, false positive; TN, true negative; FN, false negative

**Table 1.** VLMs Evaluated in This Study.

| Name of VLM | Model characteristics | Project URL |
| --- | --- | --- |
| RadVLM | A 7B multitask conversational VLM for CXR image interpretation, built on LLaVA-OneVision-7B and trained on over 1 million image–instruction pairs for report generation, abnormality classification, visual grounding, and multi-turn dialogue. | https://huggingface.co/KrauthammerLab/RadVLM-info |
| gpt-4o-mini | A commercially available multimodal model from OpenAI that accepts text and image inputs and generates text outputs. | https://developers.openai.com/api/docs/models/gpt-4o-mini |
| Qwen3-VL-8B-Instruct | A general-purpose 8B vision-language model with long-context and multimodal reasoning capabilities. | https://huggingface.co/Qwen/Qwen3-VL-8B-Instruct |
| MedGemma-4b-it | Developed by Google, MedGemma-4b-it is a 4B instruction-tuned medical VLM designed for the multimodal understanding of medical text and images, including radiology. Its training incorporates medical text, Q&A datasets, and various medical imagery. | https://huggingface.co/google/medgemma-4b-it |
| LLaVA-Rad | A lightweight 7B radiology multimodal model for CXR report generation, built on the LLaVA v1.5 architecture with BiomedCLIP-CXR and Vicuna-7B-v1.5. | https://huggingface.co/microsoft/llava-rad |
| CheXpert Plus Model | A chest X-ray findings generation baseline model trained on MIMIC-CXR and CheXpert Plus. | https://huggingface.co/IAMJB/chexpert-mimic-cxr-findings-baseline |

Abbreviation: CXR = chest radiograph; VLM = vision-language model.

The preprocessed JPEG images were resized according to the input requirements of each VLM and used as inputs to the models. The prompt for these VLMs was “Draft a concise report for this image“; however, if available, model-specific sample codes or recommended prompts on the model project page were used. This approach was adopted because the study evaluated realistic zero-shot implementations rather than a strictly prompt-controlled benchmark. The inference procedures differed across models. RadVLM and the CheXpert Plus Model were run locally using the Python transformer package, following the sample code provided on their respective Hugging Face project pages. gpt-4o-mini was accessed through the OpenAI API using the Python OpenAI package. Qwen3-VL-8B-Instruct runs on vLLM in Docker as an OpenAI-compatible server. MedGemma-4b-it was served through a FastAPI-based OpenAI-compatible server that internally used the image-text-to-text pipeline of the Python transformer package. LLaVA-Rad was executed using the GitHub source code to launch a FastAPI-based OpenAI-compatible server. For these OpenAI-compatible servers, inferences were performed by sending requests through an OpenAI API client. The textual outputs from the VLMs were saved in CSV or JSON format. The narrative reports generated using VLMs were independently reviewed by two board-certified radiologists and converted into binary labels indicating the presence or absence of pulmonary nodules. The two radiologists were blinded to model identity. Binarization was performed according to the predefined rules. Reports were classified as positive if they explicitly described a pulmonary nodule, nodular opacity, or mass-like lesion, and negative if they indicated no pulmonary abnormalities or focal lesions. Ambiguous expressions, such as “possible nodule” or “cannot exclude a nodule,” were classified as positive because they would warrant clinical attention in a detection-support setting. Discrepancies were resolved by a consensus between the two board-certified radiologists.

### Evaluation Metrics and Statistical Analysis

The VLMs were evaluated in a zero-shot setting; therefore, cross-validation (e.g., k-fold cross-validation) and dataset splitting were not performed. As shown in Figure 2, four counts of true positives (TP), true negatives (TN), false positives (FP), and false negatives (FN) were determined from the binary ground truth and binarized VLM outputs across the 247 CXR images. The following evaluation metrics were calculated:

Sensitivity (= Recall) = TP / (TP + FN)

Specificity = TN / (TN + FP)

Accuracy = (TP + TN) / (TP + TN + FP + FN)

Precision = TP / (TP + FP)

F1 =2·Precision·Recall / (Precision + Recall)

In addition to the evaluation metrics, detection performance was evaluated based on the subtlety grade assigned in the JSRT database [3,8]. For nodule-positive CXR images, the detection rate was calculated separately for each of the five subtlety grades as the number of correctly detected nodules divided by the total number of nodules in that grade. The subtlety grades were analyzed in the following order as defined by the JSRT database: Grade 5, obvious; Grade 4, relatively obvious; Grade 3, subtle; Grade 2, very subtle; and Grade 1, extremely subtle. These grade-specific detection rates were summarized descriptively and not subjected to formal statistical testing due to the limited number of cases in each grade. Differences in sensitivity, specificity, and accuracy between the models were assessed using McNemar’s tests for matched binary outcomes. For each VLM pair, the differences in each metric were compared using McNemar’s test, and the resulting 15 (= _6_C_2_) p-values were judged to be statistically significant after Holm correction for multiple comparisons [13]. Accordingly, statistical significance was determined after Holm’s correction rather than by applying an unadjusted p-value threshold of < 0.05.

## Results

All six VLMs were evaluated on 247 chest radiographs from the JSRT database: 154 lung nodule cases and 93 without. Table 2 summarizes the performance metrics of the evaluated models. Among the six VLMs, the RadVLM correctly classified 110 of 247 cases and achieved the highest accuracy (44.5%). RadVLM correctly classified all 93 nodule-negative cases as negative with a specificity of 100.0%. In addition, its precision was 100.0%, indicating that all cases judged as positive by RadVLM were truly nodule-positive. However, this highly conservative behavior was accompanied by a low sensitivity of 11.0% (17/154), indicating that most true nodules were missed. The F1 score for the RadVLM was 19.9%. These results indicate that in this zero-shot evaluation, RadVLM tended to prioritize avoiding false positives over detecting nodule-positive cases.

**Table 2.** Performance of VLMs for Lung Nodule Detection on the JSRT Database.

| Name of VLM | Sensitivity (= Recall) | Specificity | Accuracy | Precision | F1 |
| --- | --- | --- | --- | --- | --- |
| RadVLM | 11.0% (17/154) | 100.0% (93/93) | 44.5% (110/247) | 100.0% | 19.9% |
| gpt-4o-mini | 6.5% (10/154) | 93.5% (87/93) | 39.3% (97/247) | 62.5% | 11.8% |
| Qwen3-VL-8B-Instruct | 7.8% (12/154) | 88.2% (82/93) | 38.1% (94/247) | 52.2% | 13.6% |
| MedGemma-4b-it | 5.2% (8/154) | 100.0% (93/93) | 40.9% (101/247) | 100.0% | 9.9% |
| LLaVA-Rad | 27.3% (42/154) | 71.0% (66/93) | 43.7% (108/247) | 60.9% | 37.7% |
| CheXpert Plus Model | 0.6% (1/154) | 95.7% (89/93) | 36.4% (90/247) | 20.0% | 1.3% |

Footnote: Data are percentages, with numerator/denominator in parentheses where applicable.
Abbreviation: VLM = vision-language model; JSRT = Japanese Society of Radiological Technology.

The LLaVA-Rad exhibits contrasting patterns. LLaVA-Rad showed the highest sensitivity among the 6 VLMs at 27.3% (42/154) and achieved the highest F1 score (37.7%). However, this gain in sensitivity resulted in a marked decrease in specificity to 71.0% (66/93), with precision and accuracy of 60.9% and 43.7%, respectively. Thus, although LLaVA-Rad detected more nodule-positive cases than RadVLM, it produced substantially more false positive results. In contrast, RadVLM produced no false positives but detected far fewer true nodules. Accordingly, these two models represented opposite extremes of the sensitivity-specificity trade-off in the present comparison: RadVLM was the most conservative model, whereas LLaVA-Rad was the most detection-oriented.

Table 3 presents nodule detection rates stratified by subtlety grade. Detection tended to be higher for more obvious nodules, although this pattern varied across models. For grade 5 nodules, LLaVA-Rad achieved the highest detection rate, 83.3% (10/12), followed by RadVLM at 58.3% (7/12). In contrast, detection of less obvious nodules was limited; for grade 1 nodules, detection rates were 12.0% (3/25) or lower across all models. These findings indicate that zero-shot VLMs had difficulty detecting subtle and extremely subtle nodules.

**Table 3.** Detection Rates of Lung Nodules Stratified by VLM and Subtlety Grade on the JSRT Database.

| Name of VLM | Subtlety grade<br>5 | Subtlety grade<br>4 | Subtlety grade<br>3 | Subtlety grade<br>2 | Subtlety grade<br>1 |
| --- | --- | --- | --- | --- | --- |
| RadVLM | 58.3% (7/12) | 23.7% (9/38) | 2.0% (1/50) | 0.0% (0/29) | 0.0% (0/25) |
| gpt-4o-mini | 8.3% (1/12) | 7.9% (3/38) | 4.0% (2/50) | 3.5% (1/29) | 12.0% (3/25) |
| Qwen3-VL-8B-Instruct | 8.3% (1/12) | 7.9% (3/38) | 8.0% (4/50) | 10.3% (3/29) | 4.0% (1/25) |
| MedGemma-4b-it | 25.0% (3/12) | 7.9% (3/38) | 2.0% (1/50) | 3.5% (1/29) | 0.0% (0/25) |
| LLaVA-Rad | 83.3% (10/12) | 44.7% (17/38) | 20.0% (10/50) | 13.8% (4/29) | 4.0% (1/25) |
| CheXpert Plus Model | 0.0% (0/12) | 2.6% (1/38) | 0.0% (0/50) | 0.0% (0/29) | 0.0% (0/25) |

Footnote: Data are percentages, with numerator/denominator in parentheses where applicable. Total number of lung nodules is 154. Definition of subtlety in the JSRT database is as follows: 5 = Obvious, 4 = Relatively obvious, 3 = Subtle, 2 = Very subtle, and 1 = Extremely subtle.

Figures 3–5 show the results of the pairwise McNemar’s tests, which further clarify the position of RadVLM relative to that of the other models. As shown in the Methods section, statistical significance was judged using Holm correction. Regarding sensitivity, RadVLM showed no significant difference from gpt-4o-mini (raw p-value = 0.189) and Qwen3-VL-8B-Instruct (raw p-value = 0.405), suggesting that these three models are similarly limited in their ability to detect nodule-positive cases. By contrast, RadVLM’s sensitivity was higher than that of MedGemma-4b-it, with a raw p-value of 0.0352. RadVLM showed significantly lower sensitivity than LLaVA-Rad (raw p-value = 1.09 × 10^-5^), reflecting the substantially larger number of positive cases detected with LLaVA-Rad. Conversely, the sensitivity of RadVLM was significantly higher than that of the CheXpert Plus Model (raw p-value = 3.05 × 10^-5^). Overall, these results indicate that although RadVLM was not the most sensitive model, it outperformed the worst-performing model and was broadly comparable to several other models with low recall.

**Fig. 3.**
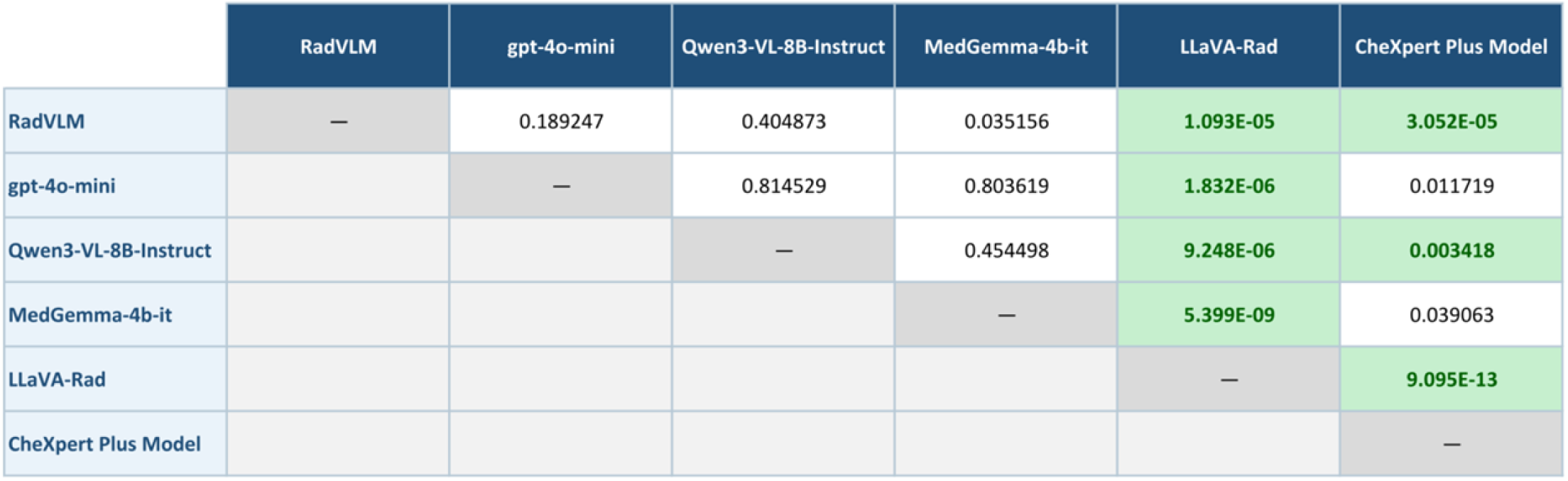
Pairwise McNemar’s test results for sensitivity among six VLMs. Cells show raw P-values for pairwise comparisons. Green cells indicate statistical significance following Holm correction for multiple comparisons. Abbreviation: VLM, vision-language model.

**Fig. 4.**
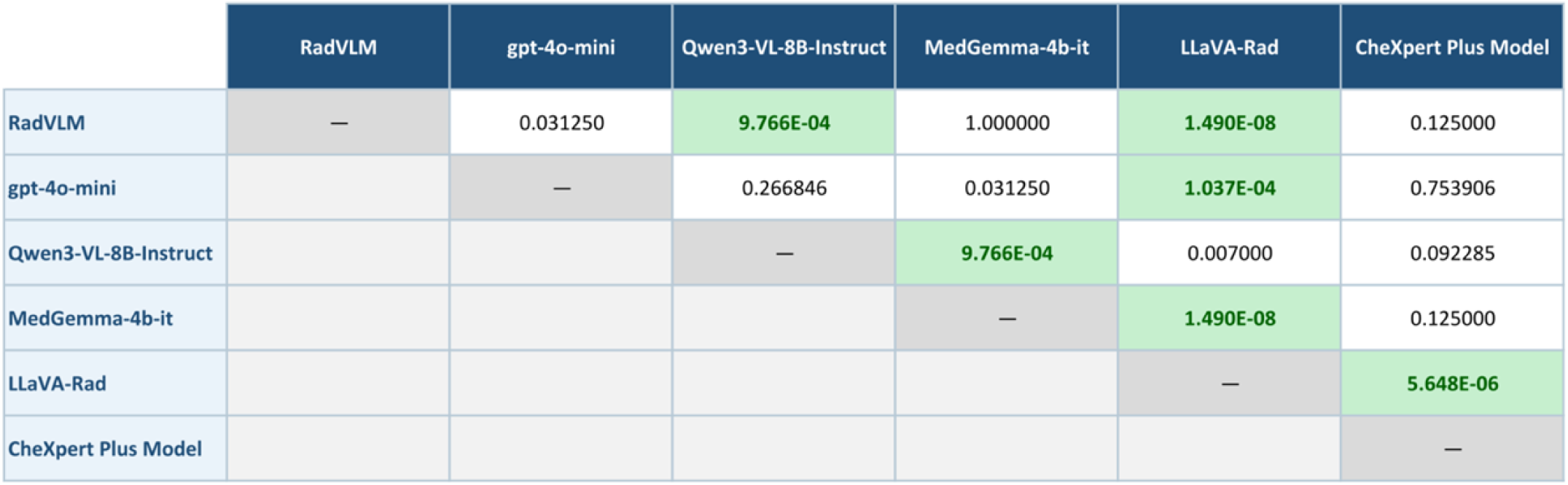
Pairwise McNemar’s test results for specificity among six VLMs. Cells show raw P-values for pairwise comparisons. Green cells indicate statistical significance following Holm correction for multiple comparisons. Abbreviation: VLM, vision-language model.

**Fig. 5.**
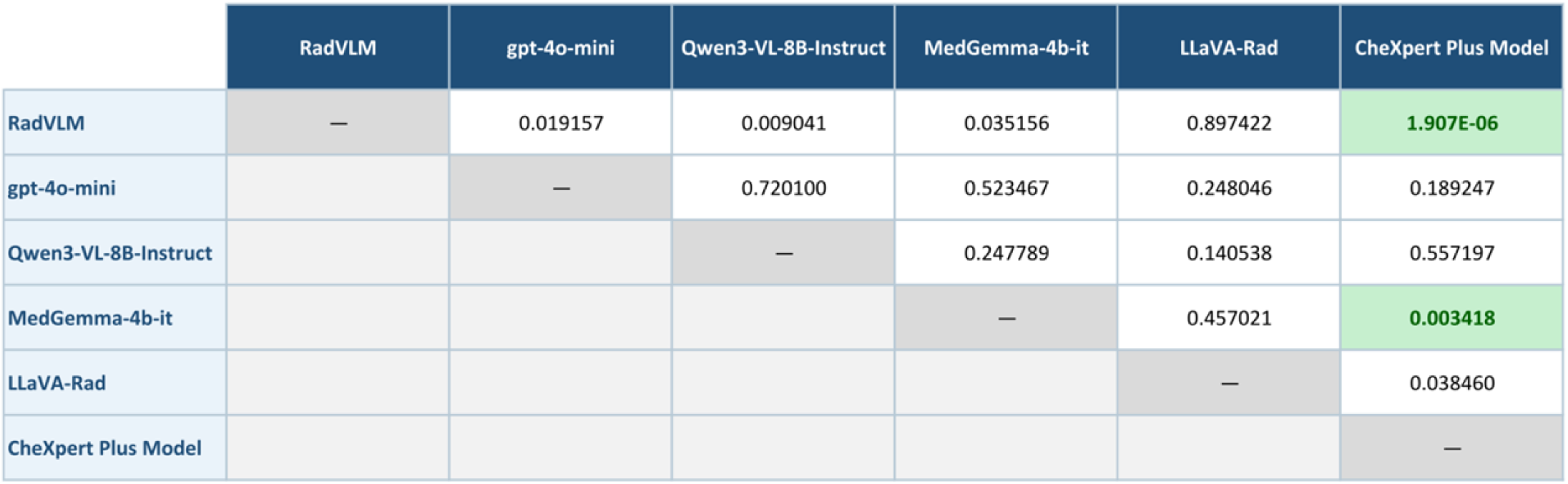
Pairwise McNemar’s test results for accuracy among six VLMs. Cells show raw P-values for pairwise comparisons. Green cells indicate statistical significance following Holm correction for multiple comparisons. Abbreviation: VLM, vision-language model.

In terms of specificity, RadVLM showed one of the best results. The specificity of RadVLM was higher than that of gpt-4o-mini (raw p-value = 0.0313) and Qwen3-VL-8B-Instruct (raw p-value = 0.000977) and far higher than that of LLaVA-Rad (raw p-value = 1.49 × 10^-8^). There was no difference between the RadVLM and MedGemma-4b-it (raw p-value = 1.0), as both models achieved 100% specificity. There was also no significant difference in specificity between the RadVLM and CheXpert Plus models (raw p-value = 0.125). These results indicate that RadVLM exhibits conservative behavior in lung nodule detection and ranks among the best-performing groups for false-positive avoidance.

Among the six models, the RadVLM exhibited the highest accuracy. The accuracy of the RadVLM was higher than that of gpt-4o-mini (raw p-value = 0.0192), Qwen3-VL-8B-Instruct (raw p-value = 0.00904), MedGemma-4b-it (raw p-value = 0.0352), and LLaVA-Rad (raw p-value = 0.897). However, the differences in the accuracy were not statistically significant. In contrast, there was a significant difference in accuracy between RadVLM and the CheXpert Plus Model (raw p-value = 1.91 × 10^-6^), despite their markedly different sensitivity and specificity profiles. These results indicate that although RadVLM and LLaVA-Rad achieved similar accuracies, the patterns of correct and incorrect classifications underlying these accuracies differed. Specifically, RadVLM maintained its accuracy with perfect specificity and high precision, whereas LLaVA-Rad compensated for its lower specificity by detecting more true-positive cases.

Overall, RadVLM showed the most conservative performance profile, with the highest accuracy and specificity, but limited sensitivity. In this zero-shot lung-nodule detection task, RadVLM was the model to achieve 100% specificity and 100% precision. Its principal weakness is the underdetection of nodules, as reflected by its low sensitivity. In contrast, LLaVA-Rad uses a detection approach that is more sensitive but significantly less specific. MedGemma-4b-it showed behavior similar to RadVLM’s in terms of conservativeness; however, its sensitivity and accuracy were lower. gpt-4o-mini and Qwen3-VL-8B-Instruct exhibited intermediate but limited overall performances, whereas the CheXpert Plus Model yielded the weakest results.

## Discussion

To the best of our knowledge, this is the first study to systematically compare the zero-shot lung nodule detection performance of six VLMs spanning general-purpose, medical-domain, and CXR-specific architectures on a standardized JSRT benchmark using a binary classification framework. A zero-shot evaluation on the JSRT database showed that multiple VLMs generally perform poorly on lung nodule detection. Among them, RadVLM demonstrated highly conservative behavior, with an accuracy of 44.5%, a specificity of 100.0%, and a precision of 100.0%, achieving the highest accuracy among the six models. However, the sensitivity was only 11.0%, suggesting that many nodule-positive cases were overlooked. In contrast, the LLaVA-Rad achieved the highest sensitivity (27.3%) and highest F1 score (37.7%); however, its specificity dropped to 71.0%, accompanied by an increase in false positives. This comparison highlights the difficulty of judging superiority among VLMs based on a single metric and demonstrates a clear trade-off between sensitivity and specificity in lung nodule detection. Grade-specific analysis further showed that detection was relatively better for obvious nodules, but remained poor for subtle and extremely subtle nodules across models.

Our results indicate that all the models demonstrated suboptimal sensitivity for stand-alone clinical use in a zero-shot setting, consistent with the role of the multimodal generative AI described in an AJR study by Hong et al. [14]. Herein, although stand-alone zero-shot performance for lung nodule detection cannot be considered sufficient, it may be worthwhile to examine its role as a second reader or as a support for drafting reports [15].

Hong et al. showed that the acceptability of AI-generated reports was high for normal cases but low for abnormal cases: for Reader 1, 91.7% in normal cases versus 52.4% in abnormal cases; and for Reader 2, 83.2% versus 37.0% [14]. Reasons for nonacceptance included false-negative findings, false-positive findings, errors in the number or location of the findings, and hallucinations. Lung nodules are small and subtle abnormalities on chest radiographs and may be more difficult to detect with free-text VLMs than with diffuse or relatively coarse abnormal patterns. The generally low sensitivity observed, together with the presence of false positives in some models, is consistent with the previously reported tendency for abnormal cases to be more difficult [14].

The JSRT database is an excellent public dataset with clearly defined digitization specifications; however, its acquisition system, image quality, and noise characteristics may differ from those of modern large-scale training datasets. The JSRT database consists of relatively classical high-resolution, high-bit-depth images with 2048 × 2048 pixels, a pixel size of 0.175 mm, and a 12-bit grayscale [3]. Such an image distribution may act as a domain shift for VLMs trained primarily on recent large-scale datasets, such as MIMIC-CXR, CheXpert, and VinDr-CXR. Accordingly, the results of this study may reflect both model-specific capability and a mismatch between the training and evaluation distributions. Conversely, evaluation under such conditions is meaningful for investigating the generalizability of lung nodule detection.

From a methodological perspective, the framework used in this study, in which free-text outputs were reduced to binary presence/absence judgments, is meaningful. Conventional VLM evaluation depends on the similarity between the generated text and the reference text and is therefore susceptible to variations in expression and redundancy. By contrast, Hong et al. reclassified AI-generated reports based on the presence or absence of tuberculosis-related abnormalities and laterality, demonstrating that a structured, binarized interpretation of free-text models is a practical approach for clinical task-based validation. The presence or absence of a lung nodule is a relatively well-suited task to such a framework and can reduce the ambiguity inherent in VLM evaluation.

Many VLMs, including RadVLM, are trained for broad purposes, such as report generation, conversation, grounding, and summarizing multiple findings; therefore, their objective functions differ from those of dedicated computer-aided diagnosis systems optimized for lung nodule detection. The model reported by Hong et al. was a generative AI system intended for comprehensive CXR report generation and combined abnormality classification with captioning. Although such model designs may be advantageous for verbalizing a broad range of abnormalities, they do not necessarily align with the high sensitivity required to detect small nodules on chest radiographs. Therefore, task-specific models may have advantages in lung nodule detection.

The preferred model depends on its intended clinical use. Specifically, high-specificity models may be advantageous when the goal is to suppress false positives or provide cautious support for report drafting, whereas in applications such as screening or support for detection, where avoiding missed lesions is more important, the current sensitivity of RadVLM is likely insufficient. The highest accuracy of RadVLM primarily results from its extremely strict handling of negative cases; it does not indicate that it can be used immediately as a stand-alone support tool for lung nodule detection.

The present evaluation was designed to reflect a realistic zero-shot implementation, rather than a strictly prompt-controlled comparison. Therefore, the observed performance differences may reflect both intrinsic model capabilities and differences in model-specific prompts, output style, and inference procedures.

This study had several limitations. First, because the textual outputs of each VLM were ultimately converted into binary labels, ROC curves based on continuous probability scores could not be calculated, and threshold selection or operating-point optimization for clinical use could not be evaluated. Second, this study assessed the performance of the stand-alone zero-shot model and did not include a reader study. Therefore, whether VLM-generated reports improve radiologist performance, reduce interpretation time, or are acceptable in clinical workflows remains to be determined. Third, our analysis was limited to the binary presence or absence of lung nodules. Performance according to nodule size, location, or benign versus malignant status was not investigated. Fourth, although the JSRT database served as an independent external benchmark for the evaluated models, only one external dataset was used. Therefore, these findings do not establish robustness across multiple datasets, institutions, acquisition systems, or contemporary clinical populations. Fifth, dedicated lung nodule detection systems or conventional computer-aided detection algorithms were not included for comparison since this study evaluated the zero-shot behavior of VLMs rather than benchmarking all available nodule detection methods. Finally, new VLMs are emerging rapidly, and future studies should include updated models as they become available [16–18].

In conclusion, our findings suggest that current VLMs show limited zero-shot generalizability for lung nodule detection in the JSRT database, with marked trade-offs between sensitivity and specificity. Their near-term value may lie more in radiologist-assisted workflows than in stand-alone detection.

## Data Availability

The CXR dataset is avaialbe from http://db.jsrt.or.jp/eng.php. Except the CXR dataset, all data produced in the present study are available upon reasonable request to the authors.

## Acknowledgments

We thank Kazuyuki Ohmura and Akie Katsuki (GE HealthCare, Japan) for their cooperation with this study.

